# Changes in rest-activity rhythms in adolescents as they age: associations with brain changes and behavior in the ABCD study

**DOI:** 10.1101/2024.03.19.24303825

**Authors:** Rui Zhang, Melanie Schwandt, Leah Vines, Nora D. Volkow

**Author notes:** ***Correspondence to*** Rui Zhang, Nora D. Volkow, Laboratory of Neuroimaging, National Institute on Alcohol Abuse and Alcoholism National Institutes of Health, 10 Center Drive, Room B2L124: MSC 1013 Bethesda, MD 20892-1013, USA.

## Abstract

**Background:** Adolescents with disrupted rest-activity rhythms (RAR) including shorter sleep duration, later sleep timing and low physical activity levels have higher risk for mental and behavioral problems. However, it remains unclear whether the same associations can be observed for within-subject changes in RAR.

**Methods:** Our longitudinal investigation on RAR used Fitbit data from the Adolescent Brain Cognitive Development (ABCD) Study at the 2-year (FL2: aged 10-13 years) and 4-year follow-up (FL4: aged 13-16 years). 963 youths had good-quality Fitbit data at both time points. In this study we examined changes in RAR from FL2 to FL4, their environmental and demographic contributors as well as brain and behavioral correlates.

**Results:** From FL2 to FL4, adolescents showed decreases in sleep duration and physical activity as well as delayed sleep timing (Cohen’s d .44-.75). The contributions of environmental and demographic factors to RAR changes were greatest to sleep timing (explained 10% variance) and least to sleep duration (explained 1% variance). Delays in sleep timing had stronger correlations with behavioral problems including greater impulsivity and poor academic performance than reductions in sleep duration or physical activity. Additionally, the various brain measures differed in their sensitivity to RAR changes. Reductions in sleep duration were associated with decreased brain functional connectivity between subcortical regions and sensorimotor and cingulo-opercular networks and with enhanced functional connectivity between sensorimotor, visual and auditory networks. Delays in sleep timing were mainly associated with grey matter changes in subcortical regions.

**Conclusions:** The current findings corroborate the role of sleep and physical activity in adolescent’s brain neurodevelopment and behavior problems. RAR might serve as biomarkers for monitoring behavioral problems in adolescents and to serve as potential therapeutic targets for mental disorders.

## INTRODUCTION

Different aspects of circadian rest-activity rhythms (RAR) including sleep timing, sleep duration and physical activity are strongly associated with mental health in adolescents. Since the launch of the Adolescent Brain Cognitive Development (ABCD) study, these associations have been replicated with this large longitudinal sample of over 10,000 adolescents, among which sleep duration has been the most studied. Adolescents with shorter sleep duration as reported by their parents at their baseline assessment showed greater internalizing problems and impulsivity cross-sectionally and longitudinally ^1–5^, which appeared to be mediated by cortico-striatal functional connectivity ^2,4^. Starting in its second year, the ABCD study implemented actigraphy to provide with objective sleep measures. A recent ABCD study applied a machine learning approach to identify brain correlates of actigraphy measures of sleep duration and confirmed the association between subcortical-parietal functional connectivity and longer sleep duration^6^. Additionally, this study also revealed an association between strengthened sensory-motor connectivity and shorter sleep duration ^6^.

In comparison, sleep timing and chronotype have been less examined in the ABCD cohort despite accumulating evidence from other national studies showing that later sleep timing and eveningness were associated with greater risk for drug use in adolescents ^7–9^. Additionally, meta-analyses have demonstrated associations of eveningness with mood-related disturbances, anxiety and a greater risk of psychotic symptoms ^10^. Interestingly, stronger and more consistent associations of mood instability and impulsivity were reported for late phase timing than for sleep duration, sleep efficiency or sleep quality as measured by actigraphy ^11^. Thus far, studies have only examined associations between evening chronotype and impulsivity using the ABCD data ^12^. Furthermore, there is growing evidence suggesting that physical activity not only lowers risks for developing internalizing problems including depression and anxiety, but also improves the symptoms in people suffering from mental health problems ^13,14^. Similar effect was not found for externalizing symptoms ^15^. In line with this, a recent ABCD study showed that adolescents with higher levels of physical activity measured by actigraphy displayed lower psychosis-like experiences and internalizing symptom severity ^16^. Finally, individual’s sleep and physical activity level appears to be strongly influenced by environmental factors and social contexts. ABCD findings have highlighted the importance of socioeconomic status, household income, marital status of parents, race and sex ^17–19^, consistent with findings from non-ABCD studies ^20^. Together, emerging evidence including from ABCD studies suggests a strong relationship between sleep duration, sleep timing, physical activity and mental health. However, the investigations have been focused on inter-individual differences in RAR and their associations with mental health including longitudinal associations between baseline RAR and mental health at follow-up and/or vice versa. So far, no studies have examined within-subject changes in objective RAR measures and their environmental, brain and behavioral correlates. In the current study, we aimed to fill this research gap by addressing three questions 1) what environmental and demographic factors contribute to individual’s RAR changes, 2) what brain measures are associated with individual’s RAR changes, 3) what behavioral changes parallel individual’s RAR changes.

## METHODS AND MATERIALS

### Participants

In the current study, data from the ABCD data release 5.1 (2024) were used. The longitudinal data includes 11,868 participants recruited from 21 sites across the United States. Informed consent from the primary caregiver and assent from the children were obtained. The study was approved by the Institutional Review Boards of all local study sites. For analyses, we selected a subsample of n=963 subjects who had valid Fitbit data at both the 2-year (FL2) and 4-year follow-ups (FL4). The days between FL2 and FL4 Fitbit measures varied from 300 to 1100 days among individuals. The selection criteria were outlined below.

### RAR assessment

Sleep and step counts were collected by wearable digital devices (Fitbit Charge series). A validation study showed adequate sensitivity of the Fitbit commercial device for measuring physical activity and sleep ^21^. We used derived daily sleep timing, sleep duration and step counts in our analyses. The following steps were conducted for quality control of the Fitbit data and for subject selection: 1) at least 60 minutes of continuous data per day, 2) the QC’d step value after all exclusions was greater than 80% of the daily reported level, 3) a minimum of 1000 steps per day, 4) a minimum of 3-hour sleep per night, 5) a minimum of 7-day physical activity and 7-night sleep data, 6) For Fitbit data at FL2, we only included data collected before the pandemic i.e., from Nov 2018 to March 2020. Fitbit data at FL4 were collected between Nov 2020 to March 2022, 6) only subjects who had data from both FL2 and FL4 were included.

### Measures of school, family and neighborhood environment

In the current study we only selected environmental factors that were measured at both FL2 and FL4. For school environment, the School Risk and Protective Factors Survey was applied to examine youth’s perceptions of the school climate and school engagement comprising three subscales: school environment, school involvement and school disengagement ^22,23^. For family environment, the youths were asked to complete the Family Conflict subscale of the Family Environment Scale ^24,25^. The Achenbach System of Empirically Based Assessment adult self-report was used to assess parents’ emotional, behavioral and drug use problems and the T-score of total problems was used in the analyses ^26^. Additionally, demographic characteristics including marital status, parental highest education and family income were considered as variables of interest. Neighborhood safety perceived by the youth was assessed by the “Safety from Crime” items from the PhenX ^27,28^. In total, there were 9 environmental variables included in the analyses.

### Daylength and its day-to-day variations

Increasing evidence suggests seasonal fluctuations in sleep, mood and brain functions influenced by light changes in the environment ^29,30^. As the number of days between FL2 and FL4 measures varied among subjects, whose data were collected at different locations and different time of the year, subjects’ RAR, mood and brain measures might be affected by light availability in their environment. In this study, we included daylength and rates of its day-to-day variations as variables of interest. We calculated daylength as the daytime plus civil twilight using R package “suncalc” for each subject using the geographic location of the 21 study sites and the date of the first day of actigraphy measure. Gain/losses of daylength were calculated by subtracting the daylength of the day prior to the first day of actigraphy measure from the daylength of the first day of actigraphy measure.

### Measures of mental and behavioral problems

Adolescents’ psychopathology and behavior problems were assessed by the Child Behavior Checklist (CBCL) reported by caregivers ^31^. It comprises two higher order factors: externalizing and internalizing problems and their t-scores were used. The Prodromal Questionnaire–Brief Child Version was used to measure subclinical prodromal psychosis risks. The Urgency, Premeditation, Perseverance, Sensation seeking and Positive Urgency (UPPS) impulsive behavior scale for children was used to measure different aspects of impulsivity ^32,33^. For youth’s grades at school, we used parent-reports and smaller scores indicated better academic performances. Substance and caffeine use in children was captured with the Youth Substance Use Interview. For drugs of abuse such as alcohol and nicotine use, the drug use in the past six months were assessed ^34^. Based on differences in drug use at FL2 vs at FL4, we categorized subjects into “Always drug users”, “Always non-drug users”, “Become drug users” and “No more drug users”. For caffeine intake, participants were asked to give the typical number of caffeinated drinks consumed per week in the past six months covering coffee, espresso, tea with caffeine, soda with caffeine, and energy drinks. Typical serving sizes were provided (coffee = 8 oz; espresso = 1 shot; tea = 8 oz, soda = 12 oz; energy drink = 5 oz or 2 oz for 5-h energy drink). In total, there were 11 mental and behavioral measures included in the analyses.

### Neuroimaging measures

Structural MRI, diffusion MRI and resting-state fMRI images were collected and processed following standardized ABCD protocols and processing pipeline ^35,36^. For each modality we used imaging data that were advised by ABCD imaging group for inclusion based on a series of imaging quality control (QC) criteria including raw QC, FreeSurfer QC, postprocessing QC, numbers of frame after censoring for rfMRI etc. For more details, please see https://wiki.abcdstudy.org/release-notes/imaging/quality-control.html. We further regressed out the effects of scanner type, mean head motion (for dMRI and rfMRI) and the number of remaining frames after motion censoring (for rfMRI only). For grey matter (GM) structure, we used 68 cortical thickness measures extracted from Desikan parcellation ^37^, and 19 subcortical grey matter volumetric measures (GMV) ^38^. Fractional anisotropy (FA) measures were used to assess microstructural tissue properties of 31 major white matter fiber tracts labelled by AtlasTrack ^39^. Resting state functional connectivity (RSFC) was calculated for pairs of regions between and within 13 cortical networks defined by the Gordon atlas ^40,41^, as well as for between each cortical network and each subcortical ROI (n=19). In total there were 338 unique functional connectivity measures. For each modality (GM, FA, RSFC) principal component analysis (PCA) was performed on the brain changes (FL4-FL2) to extract independent components that explained the variations in the brain changes from FL2 to FL4. The number of components was determined by eigenvalues greater than 1. To keep the number of principal components (PC) consistent across the three modalities, we chose the smallest number i.e., k=6.

### Statistics

Longitudinal RAR changes (FL4-FL2) in sleep duration, sleep timing and steps were the focus of this study and considered as dependent variables. To answer our first question of what environmental and demographic factors contribute to an individual’s RAR changes, best subsets regressions were implemented to identify the optimal set of potential predictors for RAR changes using R’s leap package. K-fold cross-validation (k=5) was used to evaluate all possible models for the predictors by computing the prediction error of each model. The model with lowest prediction error was selected. We then ran multiple regression models with the optimal set of predictors. Adjusted r^2^, model fit, significance and effect size of each predictor were reported. Nine environmental factors at FL2 and their longitudinal changes (FL4-FL2), three demographic characteristics (age, sex, race), days between FL2 and FL4 measures (i.e., changes in age), changes in daylength and rates of day-to-day variations were considered as potential predictors (24 potential predictors for model 1).

To answer the second question on brain correlates of RAR changes, a multiple regression model was applied for each modality separately. Extracted principal components of brain changes (k=6) were all included in the model as independent variables (Model 2). Multiple regression models were also used for the third question on behavioral correlates (n=11) of RAR changes (Model 3). For Model 2 and 3, sex, race and age at FL2 were included as covariates. False discovery rate (FDR) correction was applied for multiple testing (Model 1 &2 corrected for 3 tests and model 3 corrected for 11 tests). The threshold for statistical significance was set at p_FDR_<.05. All effect sizes were reported.

Days between Fitbit measures and onsite visit (interview date) or brain measures varied from −450 to 80 days among subjects. Since we were interested in associations with RAR, we excluded subjects whose brain and behavior measures were not close to their actigraphy measure i.e., greater than 30 days between them. Additionally, we excluded subjects with missing values. Thus, n=725 subjects remained for model 1, n=496-565 subjects for model 2 and n=786-809 subjects for model 3.

Model 1: RAR changes ~ environmental factors at FL2 +changes in environmental factors (FL4-FL2) + changes in daylength (FL4-FL2) + changes in rates of day-to-day variations in daylength (FL4-FL2) + days between FL2 and FL4 measures + sex + race + Interview age at FL2

Model 2: RAR changes ~ Brain PC1 (FL4-FL2) + Brain PC2 (FL4-FL2) + Brain PC3 (FL4-FL2) + Brain PC4 (FL4-FL2) + Brain PC5 (FL4-FL2) + Brain PC6 (FL4-FL2) + sex + race + Interview age at FL2

Model 3: RAR changes ~ changes in behavioral problems (FL4-FL2) + sex + race + Interview age at FL2

## RESULTS

### RAR at FL2 vs FL4

Compared to FL2, we observed shorter sleep duration (22.8 min less), delayed sleep timing (57 min later) and reduced physical activity (1378.7 steps less) at FL4 (Cohen’s d .44-.75) (**Table 1**). The changes in RAR from FL2 to FL4 varied between subjects (**Table 1 & Figure S1**). There were small but significant correlations between changes in sleep duration, sleep timing and physical activity (r=−.28 to −.10, all p<.002) (**Figure S2**).

**Table 1.** Demographic information and RAR at FL2 and FL4 (n=963)

| RAR | FL2<br>Mean (SD) | FL4<br>Mean (SD) | Paired t-<br>tests<br>P-value | Effect<br>size<br>(Cohen's<br>d) | FL4-FL2<br>Differences<br>(range) |
| --- | --- | --- | --- | --- | --- |
| Age | 11.98 (0.66) | 14.09 (0.71) |  |  |  |
| Sex (F) | 52.75% |  |  |  |  |
| Race/Ethnicity | White: 59.09%<br>Black: 5.82%<br>Hispanic: 20.87%<br>Asian: 3.01%<br>Other: 11.21% |  |  |  |  |
| Sleep<br>duration (min) | 452.11 (34.91) | 429.25<br>(40.22) | <.001 | .60<br>(medium) | [-167.33,<br>170.85] |
| Sleep timing | 23.01 (1.14) | 23.96 (1.35) | <.001 | .75<br>(medium) | [-5.04, 6.23] |
| Physical<br>activity<br>(Steps) | 9298.67 (2910.32) | 7919.93<br>(3202.83) | <.001 | .44<br>(small) | [-13618.92,<br>10050.64] |

### Environmental and demographic contributors to RAR changes

The best model (with lowest prediction error) for changes in sleep duration was composed of changes in parental psychopathology, changes in school environment, marital status of parents, changes in household income, household income at FL2, and days between FL2 and FL4 measures. Among these, only the marital status of parents at FL2 was significant but did not survive corrections for multiple comparisons (Standardized b= .07, p_uncorr_=.044, p_FDR_=.132, Cohen’s f^2^=.01) (**Figure 1**). The best model explained only 1% variance, though it was significant (p=.02).

**Figure 1.**
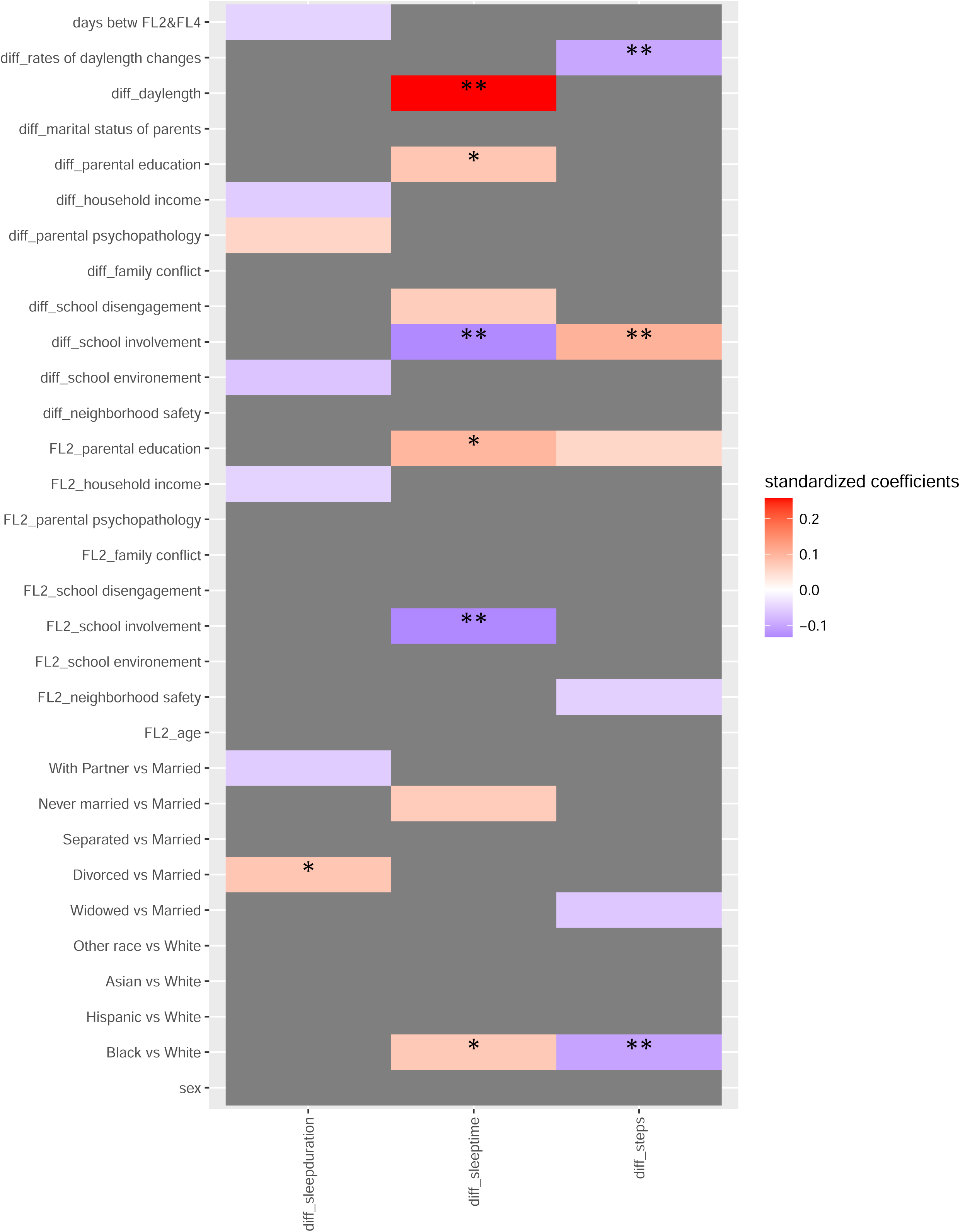
Environmental and demographic contributors to longitudinal RAR changes. Predictors included in the best model are colored. The color represents the standardized coefficients for each predictor. * denotes significant predictors for RAR changes. ** p_FDR_ <.05, * p_uncorr_<.05, diff: differences between FL2 and FL4.

For changes in sleep timing, the best model included changes in school involvement and disengagement, changes in parental education level, school involvement at FL2, parental education level at FL2, marital status at FL2, changes in daylength, and race. The model explained 10% variance and was significant (p<.001). Changes in daylength (Standardized b= .26, p_uncorr_<.001, p_FDR_<.001, Cohen’s f^2^=.07) and school involvement (Standardized b=−.13, p_uncorr_=.003, p_FDR_=.009, Cohen’s f^2^=.01) had the greatest contribution to longitudinal changes in sleep timing (**Figure 1&S3**).

For changes in physical activity, the best model comprised changes in school involvement, race, neighborhood safety at FL2, marital status of parents at FL2, parental education level at FL2 and changes in day-to-day variations of daylength. The model explained 4% variance and was significant (p<.001). Changes in school involvement (Standardized b= .10, p_uncorr_=.005, p_FDR_=.015, Cohen’s f^2^=.01), changes in day-to-day variations of daylength (Standardized b=−.10, p_uncorr_=.008, p_FDR_=.024, Cohen’s f^2^=.01) and race (black vs white: Standardized b=−.10, p_uncorr_=.005, p_FDR_=.015, Cohen’s f^2^=.01) had the greatest contribution to the changes in physical activity levels (**Figure 1&S3**).

Sex or age did not have significant effects on any RAR changes.

### Brain correlates of RAR changes

We calculated the PCs of changes in RSFC, GM and FA. For explained variance and loadings of each PC, please see **Figure 2-3** and **Figure S4-S6**. For RSFC the first 6 PCs explained 25% variance. For GMV, the first 6 PCs explained 38% variance. For FA the first 6 PCs explained 73% variance in total.

**Figure 2.**
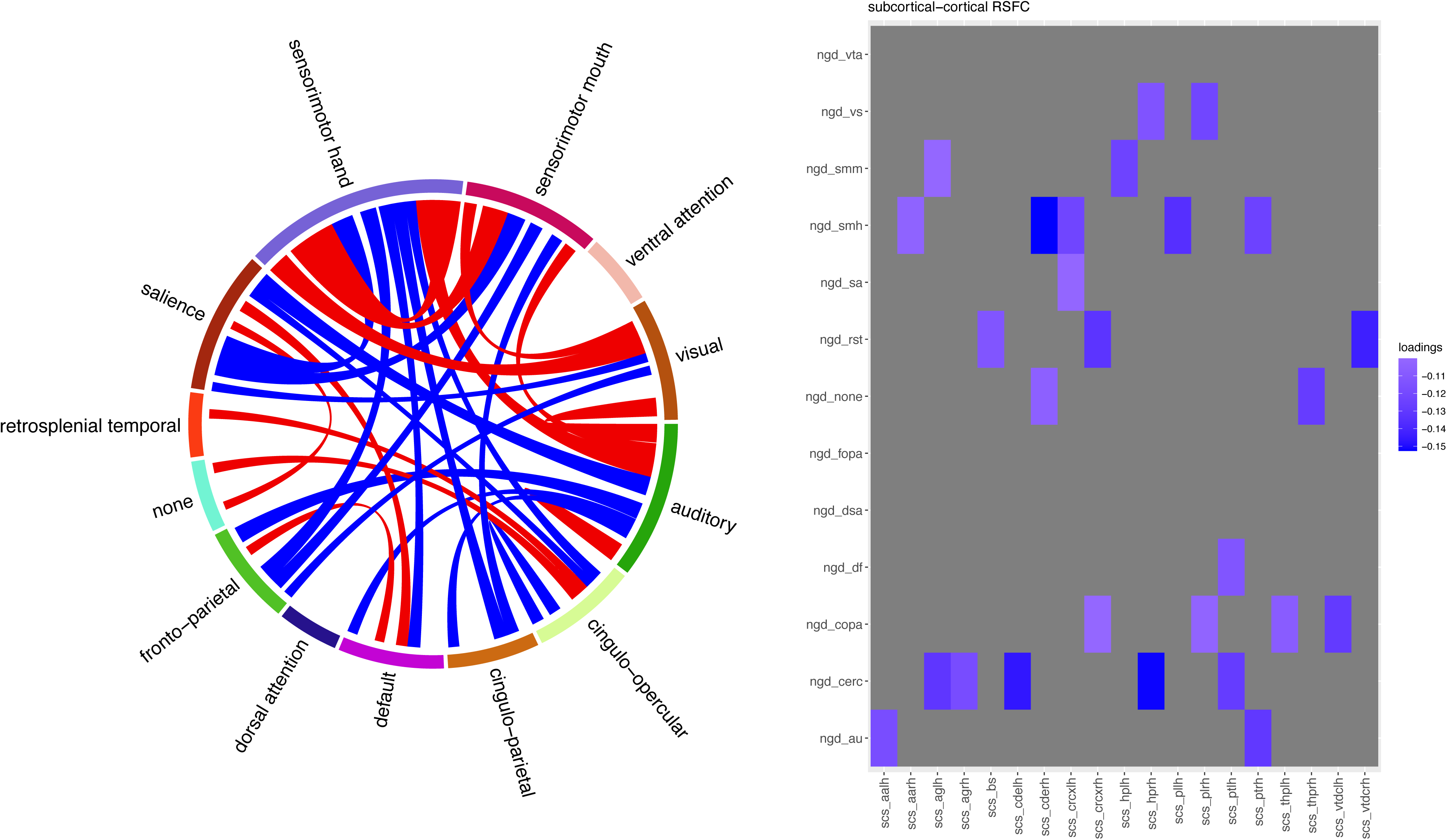
RSFC (FL4-FL2) component associated with longitudinal changes in sleep duration. Loadings of RSFC component 1 are illustrated. Left: A chord chart depicts RSFC between cortical networks. |loadings|>0.04 are illustrated. Right: RSFC between subcortical regions and cortical networks. Red indicates positive loading while blue indicates negative loadings. |loadings|>0.1 are illustrated. ngd: Gordon cortical networks, scs: subcortical regions, vta: ventral attention, vs: visual, smm: sensorimotor mouth, smh: sensorimotor hand, sa: salience, rst: retrosplenial temporal, fopa: fronto-parietal, dsa: dorsal attention, df: default mode, copa: cingulo-parietal, cerc: cingulo-opercular, au: auditory, aa: accumbens-area, ag: amygdala, bs: brain-stem, cde: caudate, crc: cerebellum, hp: hippocampus, pl: pallidum, pt: putamen, thp: thalamus-proper, vtdc: ventral diencephalon.

**Figure 3.**
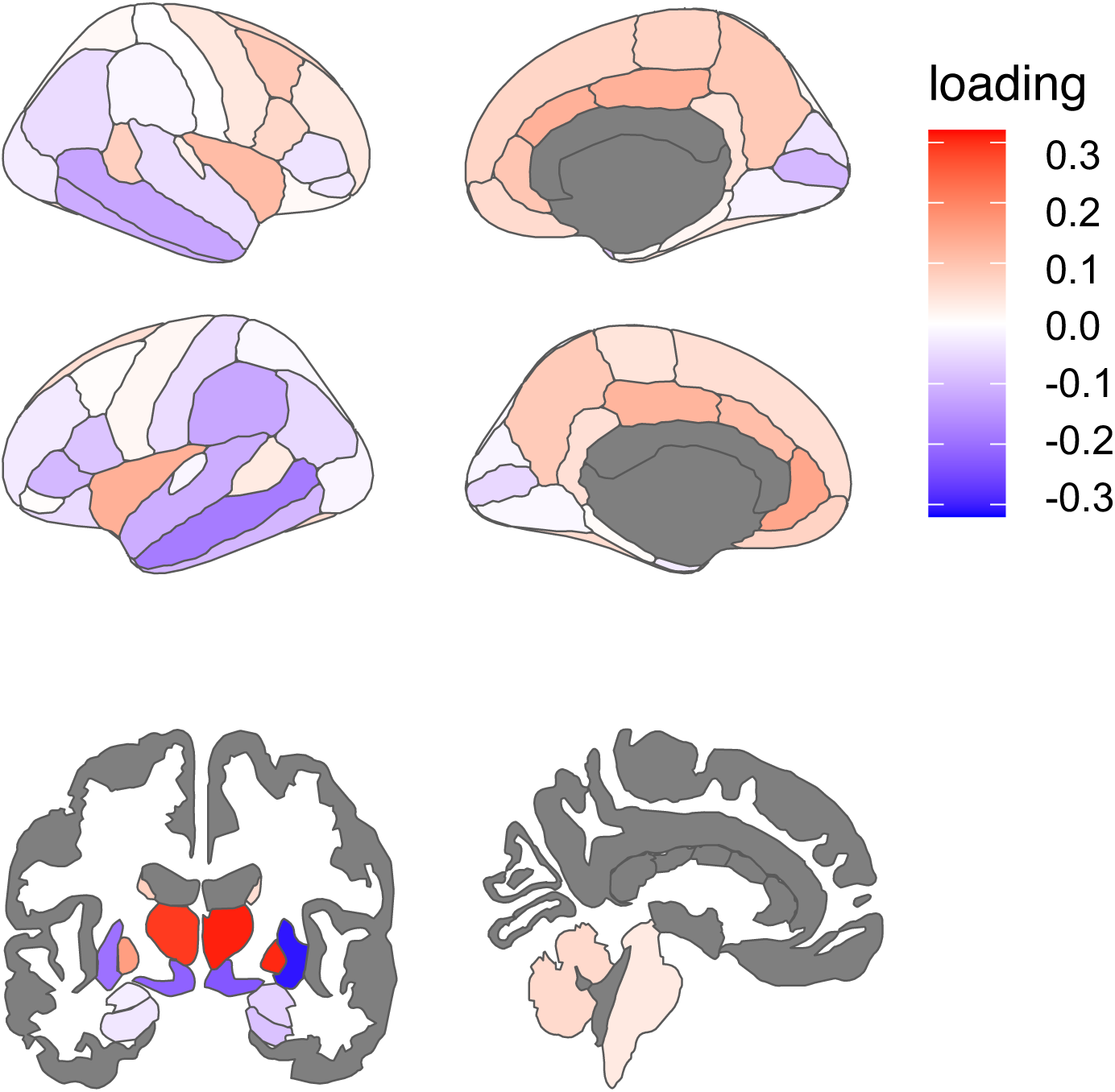
GM (FL4-FL2) component associated with longitudinal changes in sleep timing. Loadings of GM component 6 associated with changes in sleep timing are illustrated. Loadings on cortical thickness (upper), loading on subcortical grey matter volumes (bottom).

Changes in sleep duration were significantly associated with RSFC PC1, which explained 8% of variance in RSFC changes from FL2 to FL4 (Standardized b= −.14, p_uncorr_=.002, p_FDR_=.006, Cohen’s f^2^=.02). Reduced sleep duration was associated with decreased RSFC of subcortical regions with cortical networks particularly sensorimotor and cingulo-opercular networks and with increased cortico-cortical RSFC between sensorimotor, auditory, and visual networks (**Figure 2 & Table S1**).

Changes in sleep timing were significantly associated with GM PC6, which accounted for 2.8% of variance in longitudinal GM changes from FL2 to FL4 (Standardized b= .11, p_uncorr_ =.007, p_FDR_=.021, Cohen’s f^2^=.01) and was heavily loaded on subcortical regions. Delayed sleep timing was associated with subcortical GMV including decreases in putamen, accumbens, and ventral diencephalon and increases in thalamus and pallidum. Delayed sleep timing was negatively associated with cortical thickness in middle temporal regions and positively with insula and cingulate cortex (**Figure 3 & Table S2**).

Changes in physical activity were significantly associated with FA PC6, which explained 3.5% of variance in longitudinal FA changes from FL2 to FL4 but did not survive corrections for multiple comparisons (Standardized b= .09, p_uncorr_=.050, p_FDR_=.150, Cohen’s f^2^=.01). (**Table S3**).

### Mental and behavioral correlates of RAR changes

Delays in sleep timing but not reductions in sleep duration or physical activity levels, were associated with greater mental and behavioral problems including decline in school grades (Standardized b= .10, p_uncorr_=.004, p_FDR_=.031, Cohen’s f^2^=.01) and aggravation of lack of perseverance (Standardized b= .11, p_uncorr_=.001, p_FDR_=.017, Cohen’s f^2^=.01) (**Figure 4**). The associations of changes in sleep timing with psychotic risk and drug use as well as the associations of changes in physical activity with internalizing problems, academic performance and drug use did not survive corrections for multiple comparisons (**Figure 4**).

**Figure 4.**
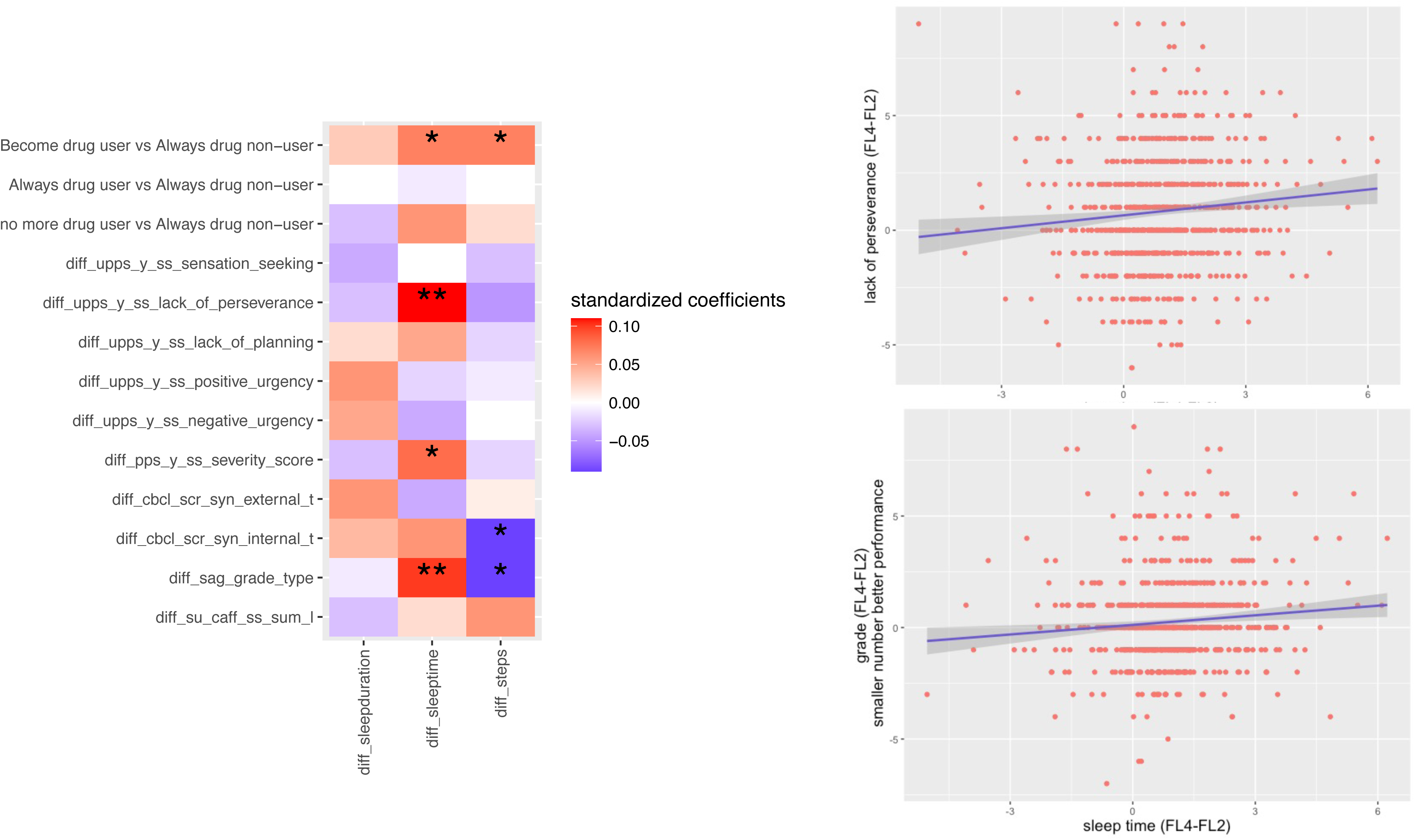
Behavioral correlates of longitudinal RAR changes. Correlations between changes in RAR and behavioral problems after regressing out sex age and race. The color in the heatmap represents the standardized coefficients. ** p_FDR_ <.05, * p_uncorr_<.05. Caff_ss_sum: weekly caffeine consumptions, pps_y_severity_score: severity score for early risks for psychosis, diff: differences between FL2 and FL4.

## DISCUSSION

The current study investigated how sleep duration, sleep timing and physical activity changed in adolescents as they grew over a 300 to 1100 day period and to identify environmental contributors to these changes as well as brain and behavioral correlates. We revealed small to medium effects of environmental factors on the RAR changes and distinct sensitivity of the various brain measures to the changes in sleep duration, sleep timing and physical activity levels within subjects. Changes in sleep timing had stronger correlations with mental and behavioral problems than the changes in sleep duration or physical activity.

From FL2 to FL4, adolescents showed decreases in sleep duration and physical activity, and delays in sleep timing. Delays in sleep timing but not reduction in sleep duration within subjects were significantly associated with decreased academic performance and increased lack of perseverance, which is in line with previous findings also based on actigraphy showing that individuals with later sleep timing had greater mood problems and impulsivity with less of an association for other sleep parameters ^11^. Compared to findings based on inter-individual differences of correlations between later sleep timing and psychosis-like symptoms ^10^, or drug use ^7–9^, in our results these correlations based on within subjects differences did not survive corrections for multiple testing. Similarly, associations between physical activity and lower internalizing problems previously reported in a cross-sectional study ^16^, did not survive corrections for multiple comparisons in our analyses based on longitudinal changes. It is possible that associations for individual differences are greatly influenced by trait and genetic factors and are thus stronger than associations for within-subject changes, which mainly reflect dynamic state changes.

We also investigated environmental and demographic contributors to the RAR changes and found important contributions from school involvement and time of the year when measures were done. Increased involvement in school including doing well in class, being part of class discussions and activities, liking school, feeling as smart as others was associated with advanced sleep timing and higher physical activity levels. Although these associations could be bi-directional, the association between school involvement at FL2 and the extent to which sleep timing was delayed from FL2 to FL4, provide support for the influence of the school environment itself on sleep timing in adolescents. Interestingly, the time of the year when measures were done had a large influence both on the changes in sleep timing and physical activity but in a different manner. Sleep timing was delayed when daylength was longer, peaking at the summer solstice; whereas physical activity level decreased when there were faster increases of daylength, peaking around the Spring equinox. However, these results must be interpreted cautiously as they might be attributed to school breaks rather than daylength itself. Furthermore, compared to white adolescents, black adolescents showed disadvantages in longitudinal RAR changes, including greater reduction in physical activity and slightly greater delays in sleep timing from FL2 to FL4. In the current analyses, among adolescents who had valid Fitbit data at both FL2 and FL4, only 5.8% were Black in contrast to 15% in the whole ABCD study sample at baseline. There might be systematic barriers that prevented good Fitbit data collection in black adolescents, which would limit our ability to investigate the effect of race on RAR and their consequences longitudinally. Future efforts to remove potential barriers are needed.

For the analyses of correlations between changes in RAR and changes in brain measures, the largest percentage of variance was obtained for the associations between sleep duration and RSFC (accounting for 8% variance of brain changes). Specifically, when an individual’s sleep duration decreased, the sensorimotor connectivity was strengthened and the subcortical functional connectivity with the sensorimotor and the salience networks was weakened. Our findings of RAR changes within individuals are consistent with those previously reported on the correlations between actigraphy-measured sleep duration and RSFC across individuals from the ABCD cohort ^6^. This indicates that these RSFCs changes not only predict differences between individuals in sleep duration but also differences in longitudinal changes within individuals. Interestingly, the observed ΔRSFC patterns associated with decreased sleep duration in adolescents had been previously identified in bereaved adults with higher anxiety levels and lower mindfulness scores ^42^. Though, in our study we did not find significant associations between changes in sleep duration and internalizing problem as reported by parents, it is possible that parent-reports, which likely reflect traits, might have not captured the state of the children at the time when the actigraphy measures were obtained. Follow up analyses of these children as they become older using self-report measures of internalizing symptoms is needed to clarify their potential association with sleep behaviors.

Changes in sleep timing were associated with change in gray matter predominantly in subcortical volumes. Previous studies in adolescents and adults ^43,44^ had reported a relationship between greater eveningness and larger GMV and cortical thickness in the medial prefrontal cortex, insula and precuneus, which are regions involved in reward evaluation and processing ^45^ and in self-referential thinking ^46^. A UK biobank study in older adults, reported that in addition to an association of eveningness with GMV in cortical regions there was also an association with subcortical regions including greater GMV in nucleus accumbens, caudate, putamen, thalamus and pallidum ^47^. We also observed positive association of delayed sleep timing with cortical thickness in reward-related regions, which has been reported by previous studies ^43,44^. In our within-subject analyses in adolescents, the strongest association with changes in sleep timing was observed for GMV changes in subcortical regions. However, unlike the reports in older adults, in the adolescents the associations with later sleep timing differed among subcortical regions such that they were associated with increased GMV in thalamus and pallidum and with decreased GMV in striatum (putamen and accumbens), and ventral diencephalon. This is consistent with findings from a cross-sectional study in children and adolescents that reported later sleep timing associated with greater GMV in pallidum ^48^. Though the pallidum is predominantly considered a motor region, a recent study reported that pallidal oscillations in local field potentials predicted sleep stages in patients with movement disorders providing evidence of its role in sleep ^49^. Associations with thalamic GMV were not surprising since the thalamus is a key region for both sleep homeostatic and circadian time keeping ^50,51^. Reduced volumes in the striatum have been linked to enhanced reward sensitivity, deficits in inhibitory control ^52^ and greater risk-taking ^53^. In line with this, greater eveningess in adolescents was associated with altered striatal reward reactivity ^54^. Our previous work has also suggested that striatal D1 receptor signaling might contribute to the association between delayed circadian rhythms and greater sensitivity to drug reward^55^.

Finally, increases in physical activity were marginally associated with greater FA in fibers from the frontolimbic pathway, though it did not survive corrections for multiple comparisons. Higher physical activity level has been reported to increase the integrity of white matter fiber tracts ^56^ and subcortical volumes in children^57^, though in youth, such associations are inconsistent^58,59^. Well-designed laboratory studies are needed to investigate how the intensity, amount and type of physical activity affect the adolescents’ brain and whether the effects vary at different developmental stages from early childhood to late adolescence.

## Conclusions

The current study identified environmental, brain and behavioral correlates of longitudinal RAR changes, offering potential interventions targets at the individual, familial and societal levels to improve sleep behaviors and physical activity in adolescents. The ongoing ABCD study provides an opportunity to follow up how RAR associations with brain and behavioral correlates changes las adolescents progress in age. With the rising popularity of wearable devices, RAR might serve as biomarkers for monitoring or as targets for treating mental and behavioral problems in adolescents. In parallel, well-designed experimental or intervention studies could help elucidate the causality behind the observed longitudinal associations between RAR, environment, brain and health outcomes.

## Disclosure

All authors declare no financial interests or potential conflicts of interest

## Data Availability

The ABCD data used in this report came from https://nda.nih.gov/study.html?id=2313.

## ABCD Acknowledgements

Data used in the preparation of this article were obtained from the Adolescent Brain Cognitive Development (ABCD) Study (https://abcdstudy.org), held in the NIMH Data Archive (NDA). This is a multisite, longitudinal study designed to recruit more than 10,000 children age 9–10 and follow them over 10 years into early adulthood. The ABCD Study is supported by the National Institutes of Health and additional federal partners under award numbers U01DA041022, U01DA041025, U01DA041028, U01DA041048, U01DA041089, U01DA041093, U01DA041106, U01DA041117, U01DA041120, U01DA041134, U01DA041148, U01DA041156, U01DA041174, U24DA041123, and U24DA041147. A full list of supporters is available at https://abcdstudy.org/nih-collaborators. A listing of participating sites and a complete listing of the study investigators can be found at https://abcdstudy.org/principal-investigators.html. ABCD consortium investigators designed and implemented the study and/or provided data but did not necessarily participate in analysis or writing of this report. This manuscript reflects the views of the authors and may not reflect the opinions or views of the NIH or ABCD consortium investigators. The ABCD data repository grows and changes over time. The ABCD data used in this report came from https://nda.nih.gov/study.html?id=2313.

