## Supplement for "Changes in rest-activity rhythms in adolescents as they age: associations with brain changes and behavior in the ABCD study"

**Figure S1** RAR at FL2 and FL4

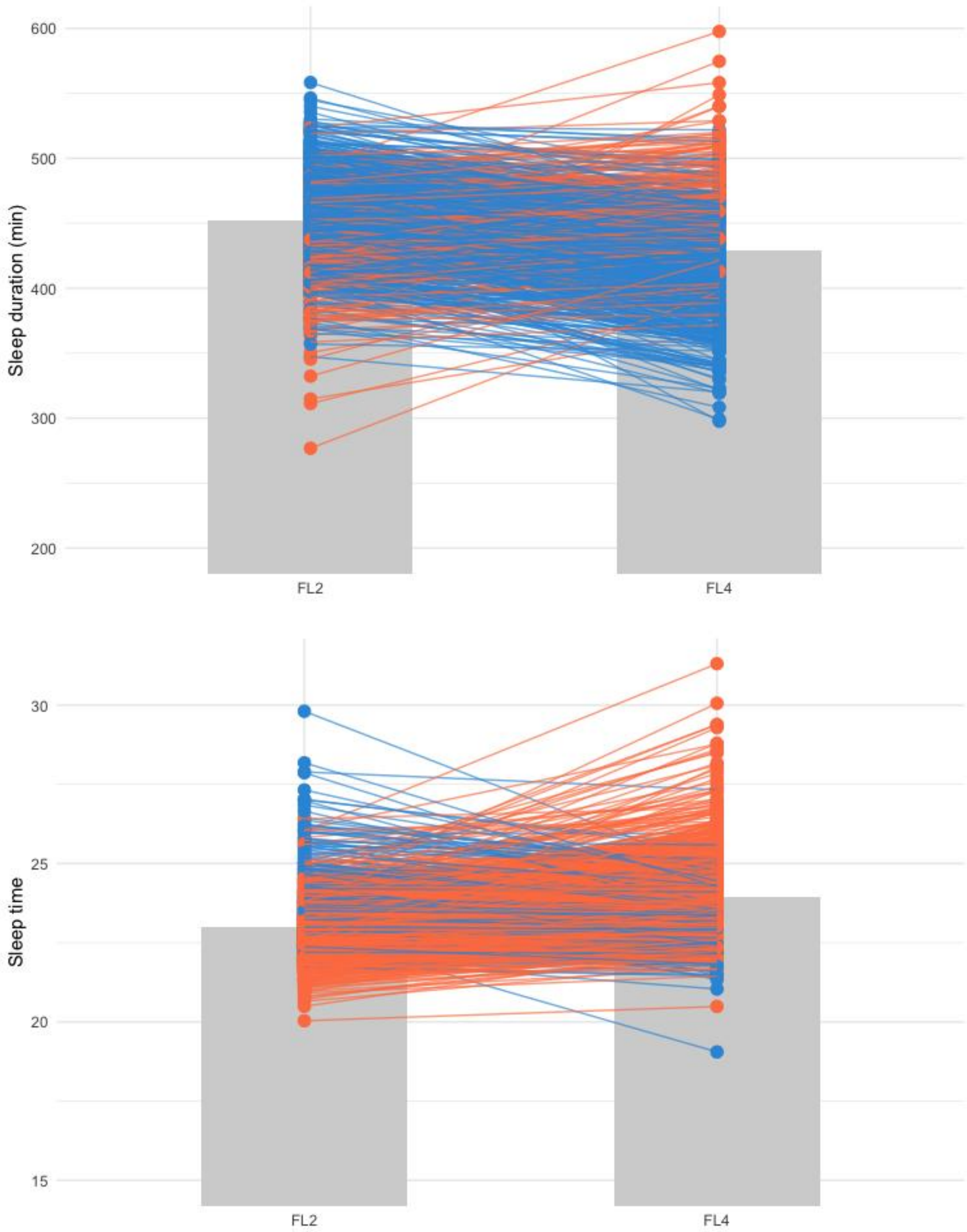

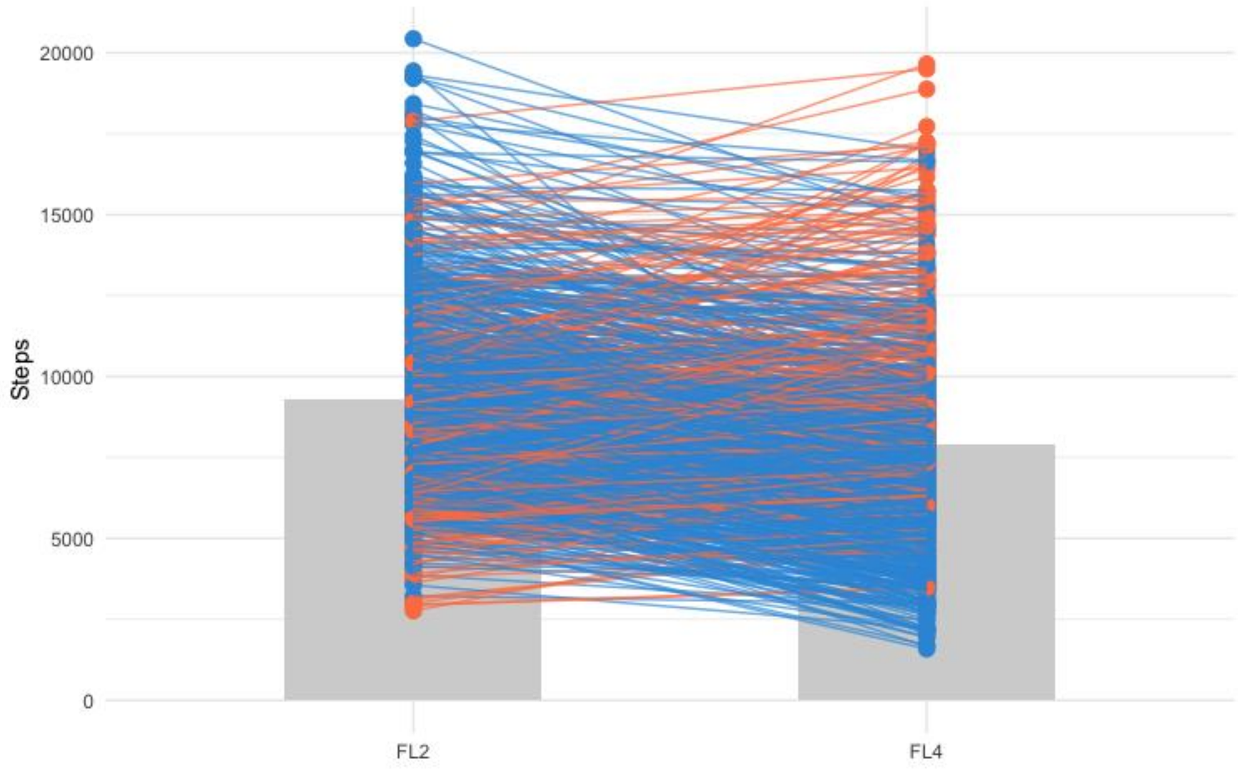

20000
15000
10000
5000
0
Steps
FL2
FL4

**Figure S2** Correlations between changes in sleep duration, sleep timing and steps (FL4-FL2)

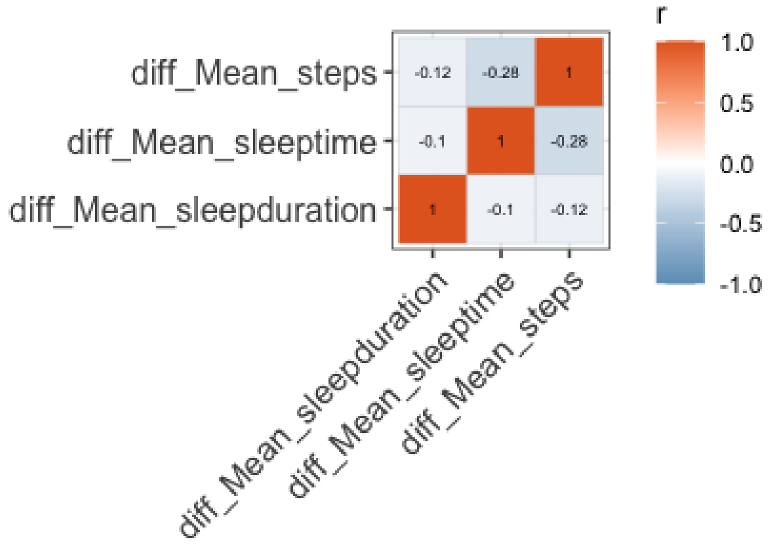

### Figure S3 Correlations between environmental contributors and RAR changes

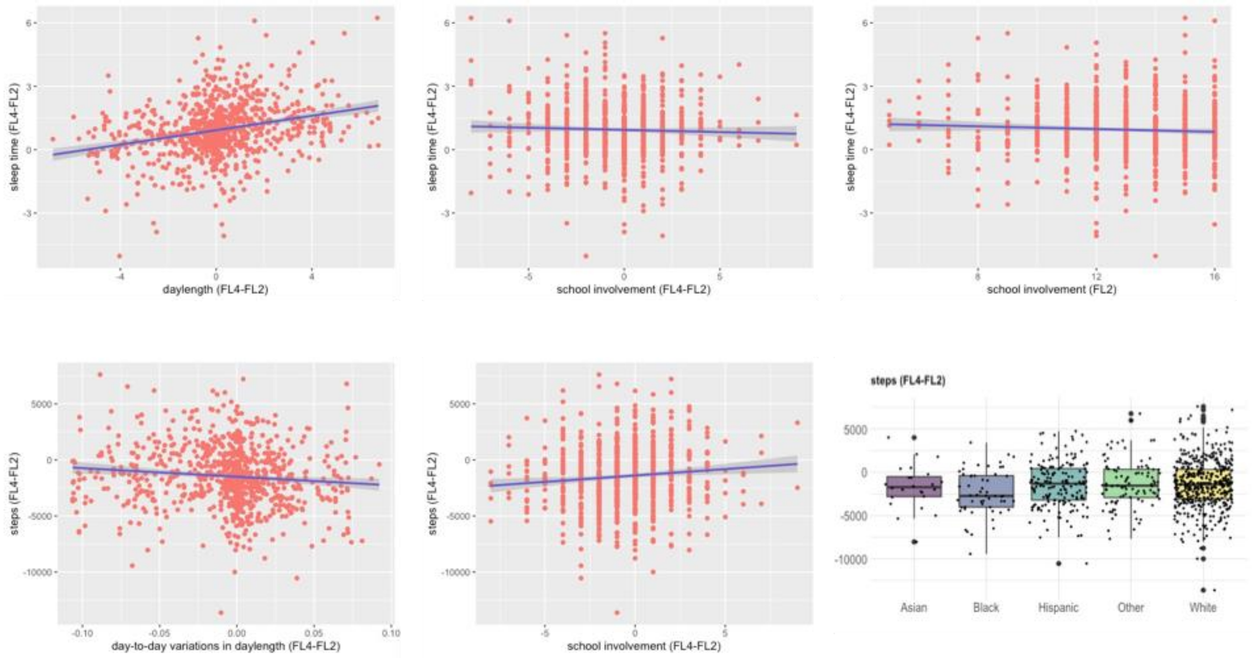

#### Figure S4 PCs of RSFC (FL4-FL2)

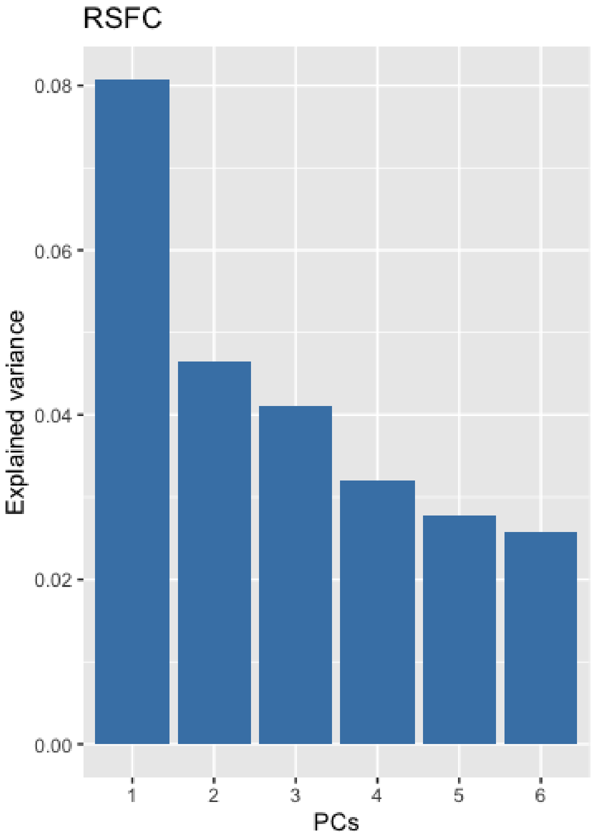


#### See Figure 2 for PC1 associated with changes in sleep duration

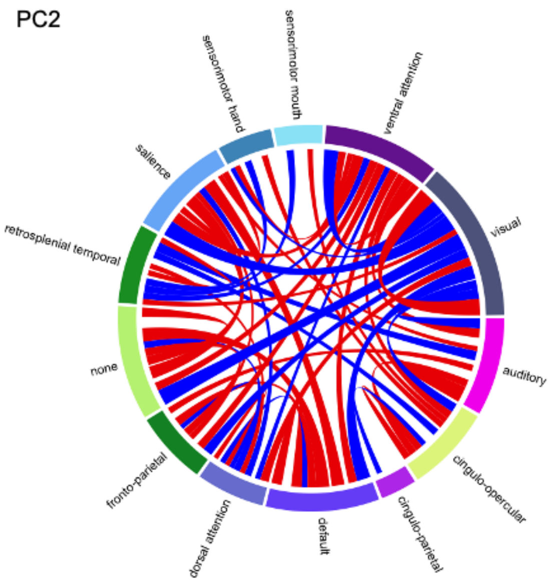


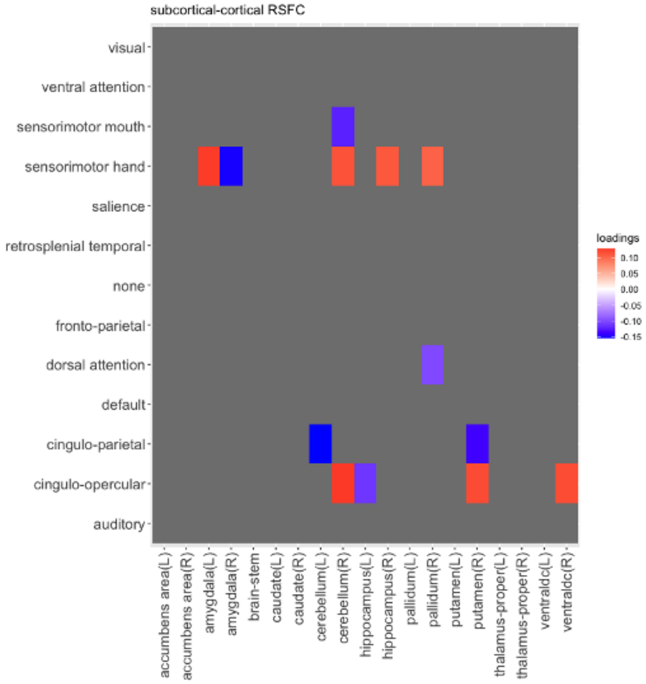

PC3

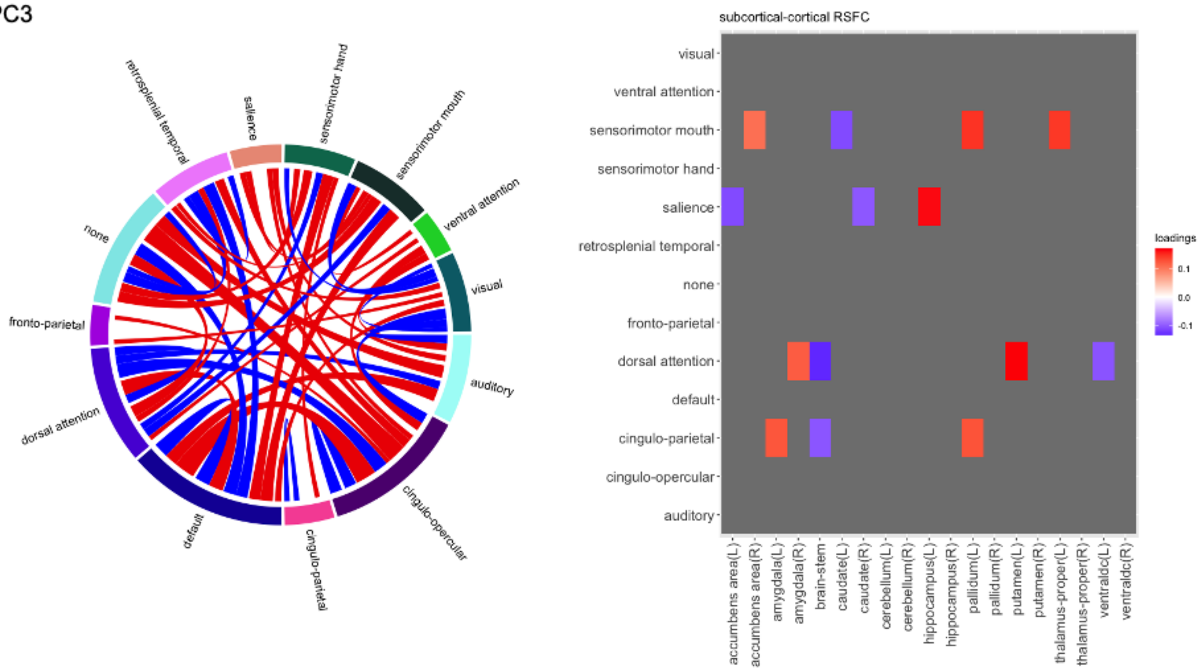
retrosplenial temporal
salience
sensorimotor hand
sensorimotor mouth
ventral attention
visual
auditory
cingulo-opercular
cingulo-parietal
default
dorsal attention
fronto-parietal
none
subcortical-cortical RSFC
visual
ventral attention
sensorimotor mouth
sensorimotor hand
salience
retrosplenial temporal
none
fronto-parietal
dorsal attention
default
cingulo-parietal
cingulo-opercular
auditory
accumbens area(L)
accumbens area(R)
amygdala(L)
amygdala(R)
brain-stem
caudate(L)
caudate(R)
cerebellum(L)
cerebellum(R)
hippocampus(L)
hippocampus(R)
pallidum(L)
pallidum(R)
putamen(L)
putamen(R)
thalamus-proper(L)
thalamus-proper(R)
ventraldc(L)
ventraldc(R)
loadings
0.1
0.0
-0.1

PC4

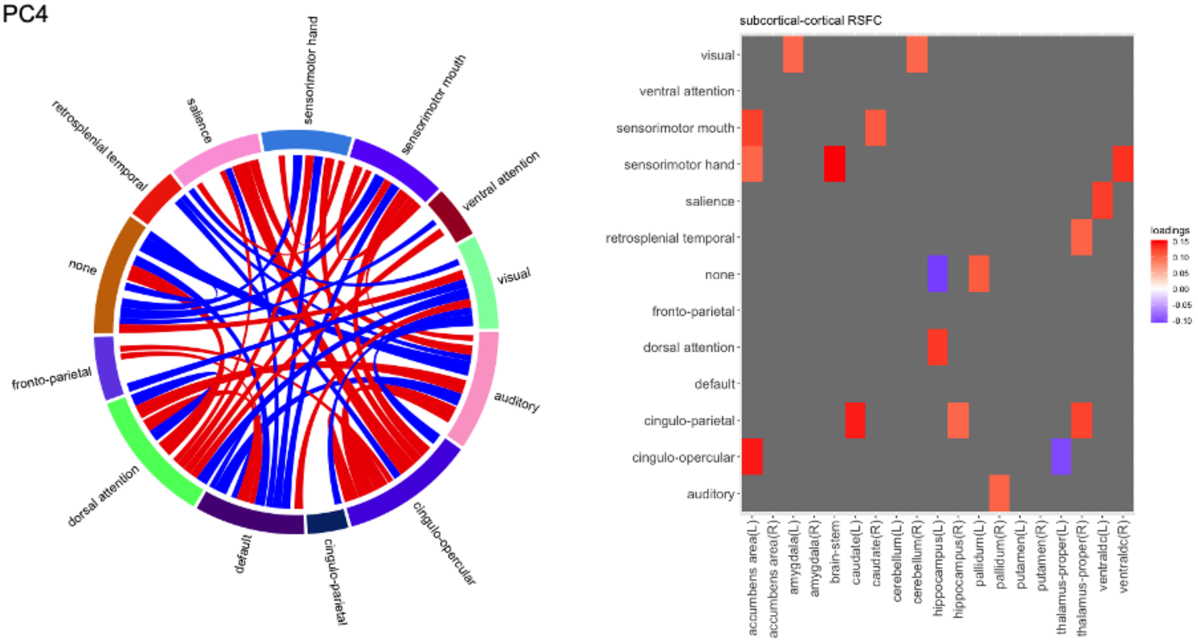
retrosplenial temporal
salience
sensorimotor hand
sensorimotor mouth
ventral attention
visual
auditory
cingulo-opercular
cingulo-parietal
default
dorsal attention
fronto-parietal
none
subcortical-cortical RSFC
visual
ventral attention
sensorimotor mouth
sensorimotor hand
salience
retrosplenial temporal
none
fronto-parietal
dorsal attention
default
cingulo-parietal
cingulo-opercular
auditory
accumbens area(L)
accumbens area(R)
amygdala(L)
amygdala(R)
brain-stem
caudate(L)
caudate(R)
cerebellum(L)
cerebellum(R)
hippocampus(L)
hippocampus(R)
pallidum(L)
pallidum(R)
putamen(L)
putamen(R)
thalamus-proper(L)
thalamus-proper(R)
ventraldc(L)
ventraldc(R)
loadings

PC5

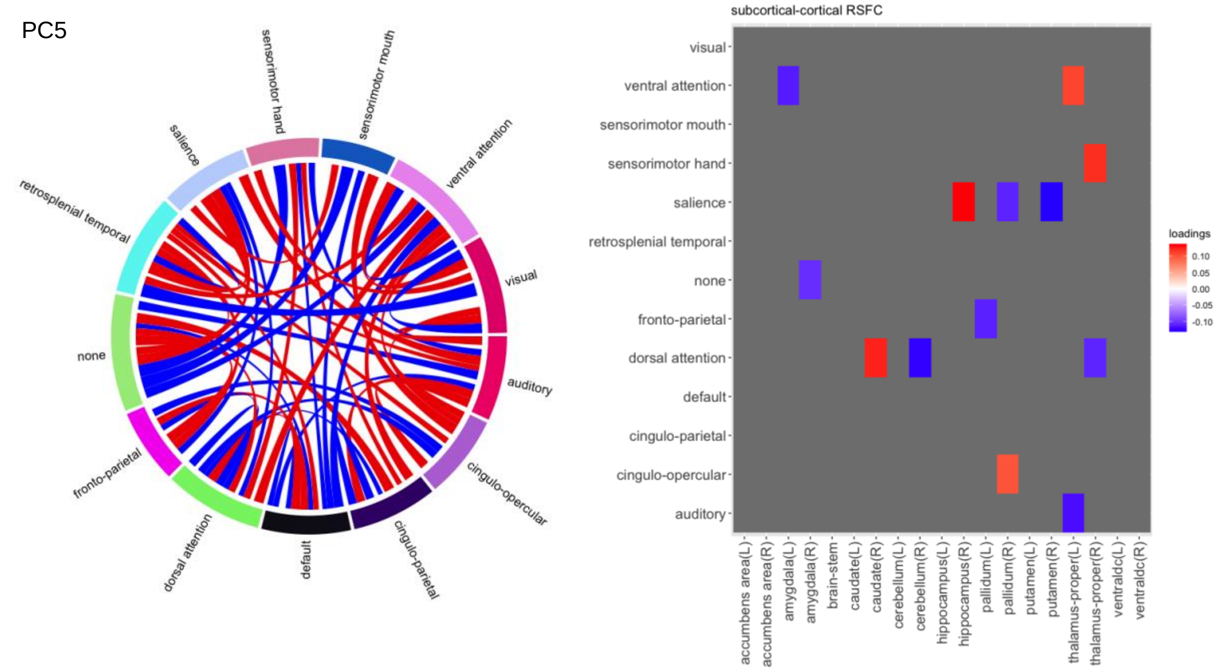
sensorimotor hand
sensorimotor mouth
ventral attention
visual
auditory
cingulo-opercular
cingulo-parietal
default
dorsal attention
fronto-parietal
none
retrosplenial temporal
salience
subcortical-cortical RSFC
visual
ventral attention
sensorimotor mouth
sensorimotor hand
salience
retrosplenial temporal
none
fronto-parietal
dorsal attention
default
cingulo-parietal
cingulo-opercular
auditory
accumbens area(L)
accumbens area(R)
amygdala(L)
amygdala(R)
brain-stem
caudate(L)
caudate(R)
cerebellum(L)
cerebellum(R)
hippocampus(L)
hippocampus(R)
pallidum(L)
pallidum(R)
putamen(L)
putamen(R)
thalamus-proper(L)
thalamus-proper(R)
ventraldc(L)
ventraldc(R)
loadings
0.10
0.05
0.00
-0.05
-0.10

PC6

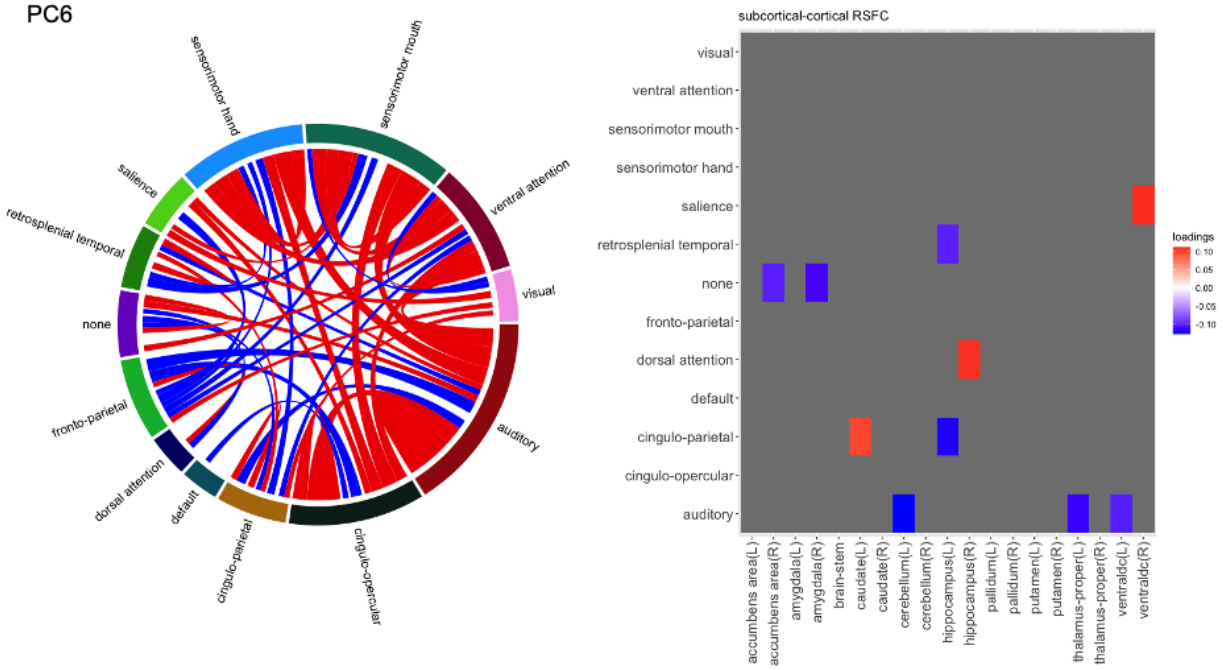
sensorimotor hand
sensorimotor mouth
ventral attention
visual
auditory
cingulo-opercular
cingulo-parietal
default
dorsal attention
fronto-parietal
none
retrosplenial temporal
salience
subcortical-cortical RSFC
visual
ventral attention
sensorimotor mouth
sensorimotor hand
salience
retrosplenial temporal
none
fronto-parietal
dorsal attention
default
cingulo-parietal
cingulo-opercular
auditory
accumbens area(L)
accumbens area(R)
amygdala(L)
amygdala(R)
brain-stem
caudate(L)
caudate(R)
cerebellum(L)
cerebellum(R)
hippocampus(L)
hippocampus(R)
pallidum(L)
pallidum(R)
putamen(L)
putamen(R)
thalamus-proper(L)
thalamus-proper(R)
ventraldc(L)
ventraldc(R)
loadings
0.10
0.05
0.00
-0.05
-0.10

**Figure S5 PCs of GM (FL4-FL2)**

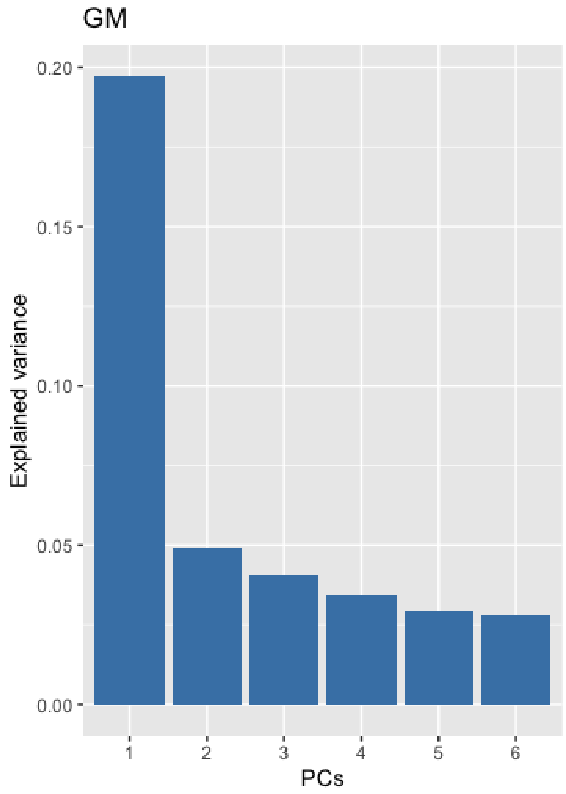

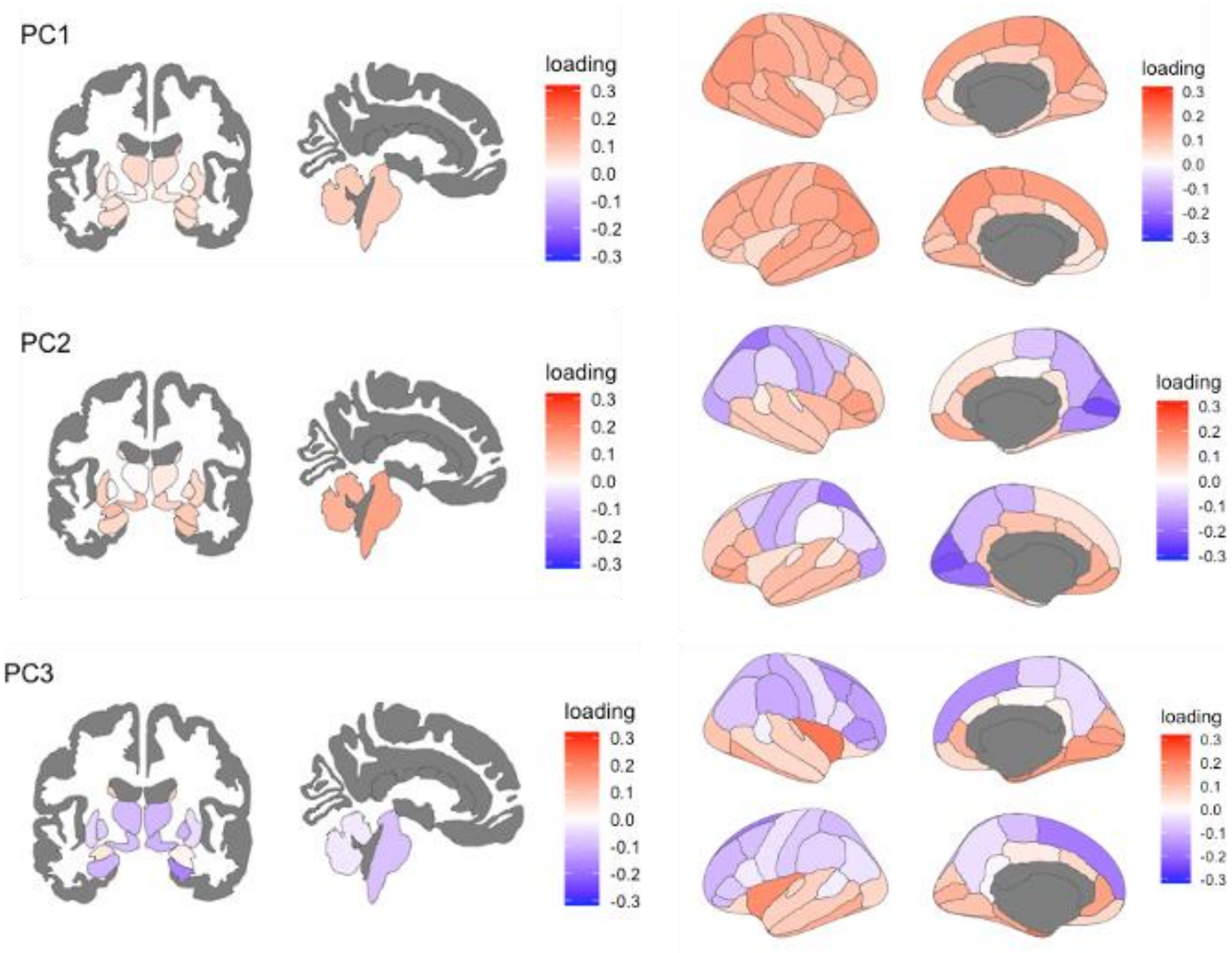

PC1
loading
0.3
0.2
0.1
0.0
-0.1
-0.2
-0.3
PC2
loading
0.3
0.2
0.1
0.0
-0.1
-0.2
-0.3
PC3
loading
0.3
0.2
0.1
0.0
-0.1
-0.2
-0.3

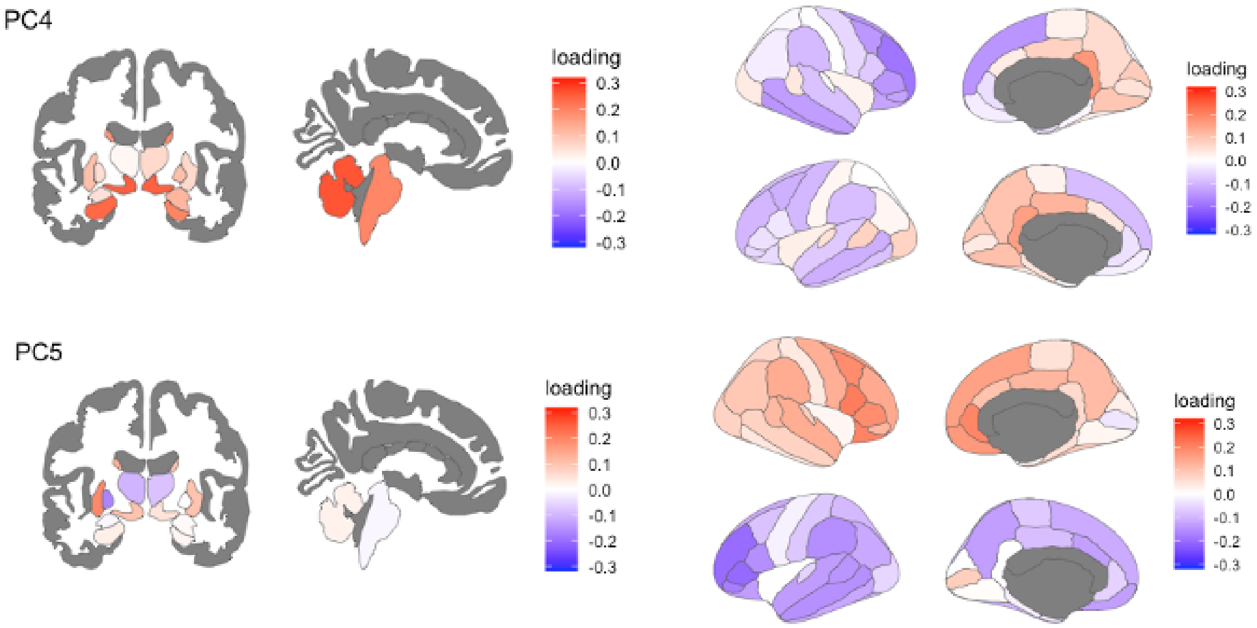


**See Figure 3 for PC6 associated with changes in sleep timing**

**Figure S6 PCs of FA (FL4-FL2)**

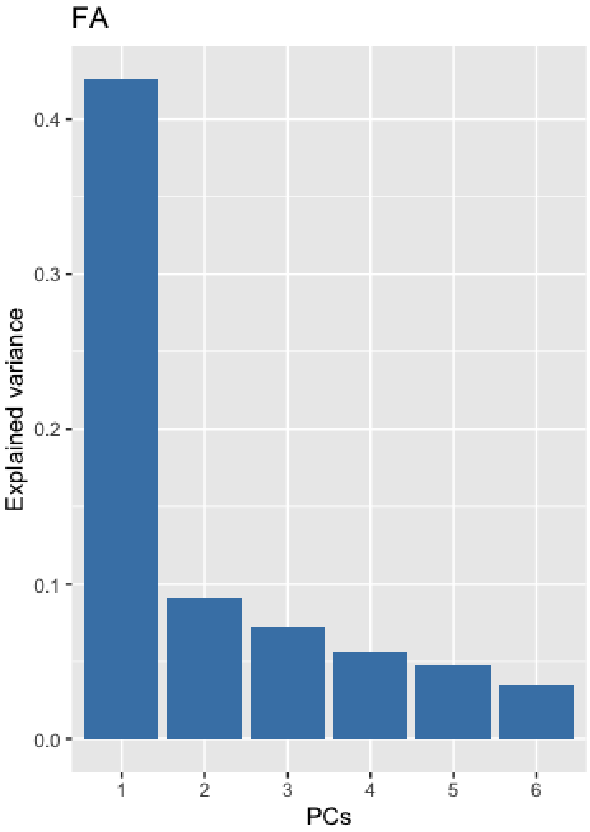


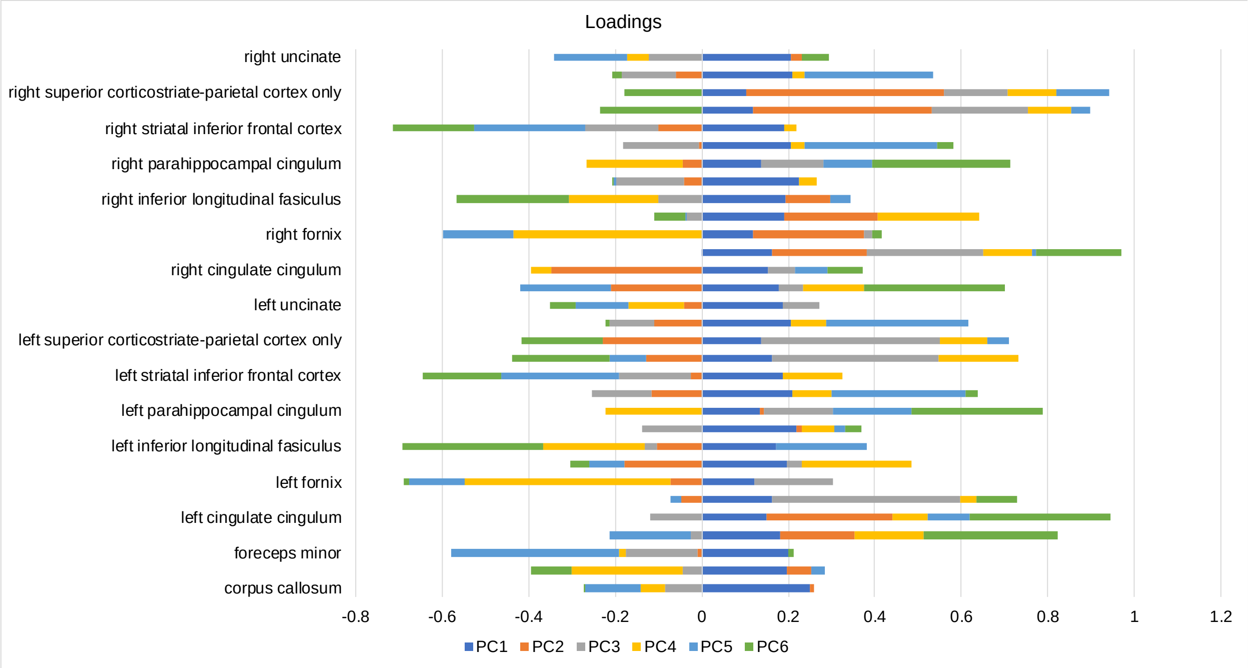
